# Engaging Community and Healthcare Stakeholders in the Design of HIV Retesting Messages: Findings from Human-Centered Design Workshops in Kenya and Uganda

**DOI:** 10.64898/2026.04.28.26351157

**Authors:** Megan A. Rabin, Alison M. Buttenheim, Kara Marson, Sabina Ogachi, Ronald Kisitu, James Ayieko, Jane Kabami, Moses R. Kamya, Shalin Desai, Kriti Chouhan, Gabriel Chamie, Harsha Thirumurthy

## Abstract

Frequent HIV testing, or “retesting,” the practice of regular HIV testing following a negative test result, among persons at high risk of HIV exposure is critical for initiating treatment early among newly infected persons and reducing the risk of HIV transmission. However, barriers to HIV retesting, such as fear of stigma, underestimating risk after a prior negative HIV test, and navigating the logistics of accessing an HIV test, have contributed to lower-than-desired retesting rates in Sub-Saharan Africa, where median time from infection to diagnosis is over 2.5 years. The Innovative Behavioral Intervention Strategies (IBIS) study aims to encourage re-testing by utilizing principles of behavioral economics and human-centered-design in a many-arm randomized trial (known as a “megatrial”) of avatar-delivered video-based messages and text messages to promote HIV retesting. In 2025, we conducted two-day focus groups in Kenya and Uganda to prototype the messages among community members and healthcare workers. An expert team engaged participants in various activities and discussions to elicit their feedback, where they reflected on factors such as local relevance, clarity, and visual appeal for each prototype. Key changes as a result of workshop feedback include standardized greetings for each arm, clearer language and refined translations, SMS language which protects participant privacy, and avatar updates for local acceptability, while maintaining core behavioral theory. The workshops generated important insights that shaped the final avatars, scripts, and messages encouraging HIV retesting which will be incorporated in the eventual trial. This study demonstrates the value of engaging end-users early in the intervention development process, and gives insight into the application of artificial intelligence (AI) to improve health behaviors in resource-limited settings.

## Introduction

Identifying simple and low-cost ways to promote behavior change has been the focus of substantial research in the social sciences and public health. In the field of behavioral economics, “nudges” have been recognized as a way to overcome people’s reluctance to engage in a wide range of behaviors that are influenced by various barriers and biases in human decision-making. Nudges such as small financial incentives, strategically framed information and messages, and changes in “choice architecture” (e.g., modifying default settings) have proven effective in promoting many health behaviors including vaccination, smoking cessation, gym attendance, and HIV testing (1–6). An important lesson from many studies of nudges and other behavioral interventions, however, is that they must be adapted to local settings and designed with end-user input in order to be effective. The field of human-centered design (HCD), which engages end-users and community members as active partners in developing solutions to problems, is well-poised to meet this need (7). Drawing on iterative prototyping and deep engagement with communities, HCD has increasingly been used in public health research and practice to ensure interventions are both acceptable and impactful (8,9).

There are a number of examples of HCD being used to design behavioral interventions that address public health priorities. For example, to address challenges in HIV prevention and treatment, one study in South Africa engaged men from communities in South Africa to help develop messages that communicate “Undetectable Equals Untransmittable,” or “U=U,” the concept that people with HIV can eliminate their risk of HIV transmission through antiretroviral treatment (ART) (10). Participants developed simple messages in their local language that addressed fears about HIV and attested to the benefits of ART. HCD was also used to co-design interventions to mitigate harm from the restrictive cultural practice of *chhaupadi,* or menstrual seclusion, in far-West Nepal (11). In this intervention development workshop, a group of women in the community were enlisted to share insights about their personal experiences and feelings concerning *chhaupadi,* then brainstorm and develop locally relevant intervention proposals.

A timely application of human-centered design and behavioral science pertains to HIV retesting, the practice of routinely testing for HIV after a prior negative test. Frequent HIV testing in persons at increased HIV risk is crucial for early diagnosis and prompt ART initiation – key steps to reducing morbidity and eliminating transmission. It can also support ongoing efforts to scale-up the use of pre-exposure prophylaxis (PrEP). The World Health Organization (WHO) recommends HIV testing every 3-6 months for adults at higher risk (12). However, the reality is that HIV retesting rates are low in Sub-Saharan Africa (SSA), and median time to HIV diagnosis after infection is more than 2.5 years (13). In a study of adults at increased risk of HIV in southwestern Uganda recruited from bars, sites of commercial sex work and transport hubs, we found that most adults (>80%) reported ever testing for HIV. However, far fewer (18%) reported regular retesting for HIV every three months in accordance with Ugandan guidelines (14).

Similar findings have been reported elsewhere in SSA, suggesting that retesting is a distinct behavior from one-time testing (15–17). These late diagnoses result in missed opportunities for HIV treatment as prevention along with avoidable morbidity and mortality.

There are various barriers to HIV testing generally. Many individuals cite fear of stigma or discrimination in their community if they test HIV positive, especially if they are part of key populations such as men who have sex with men or female sex workers (12,18,19). There are also logistical difficulties in accessing HIV testing services, such as the distance to these services, or the opportunity costs or leaving work and household obligations to visit a clinic (12). The field of behavioral economics offers additional insight into retesting rates, and why they are reinforced by prior negative test results. One such study of young women at high HIV risk in South Africa found that prior HIV testing was a significant predictor of lower perceived HIV risk (20). Another barrier to retesting is inaccurate mental models of HIV infection; for example, individuals may believe that retesting is unnecessary if a person feels healthy (21,22). Furthermore, retesting rates are curbed by decreased salience of HIV prevention over time, leading to outcomes like prevention fatigue (23,24).

In order to improve HIV retesting outcomes, it is necessary to engage end-users with services they want to use, and which are accessible to them. Yet, to date, there have been few evidence-based interventions to promote retesting among high-risk persons; a recent systematic review of interventions to promote retesting found ten studies, only three of which were in SSA (25). Prior to launching the IBIS study, a large-scale trial of video-based messages and text messages to promote HIV retesting among high-risk individuals, we generated a series of messages based on crowdsourced ideas from behavioral science experts and then used HCD to develop the messages. We conducted two-day HCD workshops with community members and healthcare workers (HCWs) in Kenya and Uganda to elicit feedback and fine-tune avatar messengers who delivered a short script for each intervention arm of the trial, as well as accompanying SMS messages. Here we describe the implementation of the human-centered design workshops and assess how participant input informed the development and adaptation of messages. This study is one of the first to develop messages that specifically promote the behavior of HIV retesting and that will be delivered in videos by artificial intelligence (AI)-generated avatars. The HCD workshops provide novel insights into the integration of automated technology such as AI avatars into health interventions in resource-limited settings, while illuminating the role of community feedback on intervention adaptation.

## Methods

Building on prior work that identified barriers to HIV retesting, we first selected candidate interventions that have proven effective in addressing similar barriers to either HIV testing or other health behaviors. We also identified and contacted an affiliate group of experts in behavioral economics and HIV testing, and invited them to suggest intervention messaging ideas for promoting HIV retesting among persons at increased risk of HIV. We then reviewed all intervention ideas that were suggested by the experts, consolidated overlapping ideas and removed those which were not feasible.

While developing the interventions, we also considered the best way to present these messages to persons engaging in HIV testing services (HTS) in Kenya and Uganda. Early ideas for delivering the intervention included scripts for counselors to read, or pre-recorded videos of counselors reading the scripts, but we decided to use avatars to ensure consistent message delivery across parent trial sites and to ease the burden of work for counselors in high-volume, HTS settings. We developed an initial script for each intervention message to be delivered in the clinic setting during post-test counseling (following a baseline, negative HIV test), and two SMS follow-up messages to be delivered at eight and eleven weeks after baseline testing to reinforce the nudge and remind participants about the importance of retesting for HIV 3-6 months after their baseline test.

We presented the intervention to a consulting team from Busara, a nonprofit organization in Nairobi, Kenya that applies behavioral science to address challenges in health and other sectors in LMICs (26). We went through several iterations of initial prototypes with Busara as they revised the nudges and we provided feedback until we were mutually satisfied with the messages. Busara contractors then translated the messages into the study languages, English, Swahili, Luo, Luganda, and Runyonkole, which local team members reviewed and revised as needed.

We worked with Consilient Talk, an organization within the research firm Consilient that conducts local, on-the-ground research for clients largely within the international development and aid space (27,28). Consilient Talk offers a suite of avatars and language tools intended for use in humanitarian and development organizations in SSA (29). The local study teams from each country chose initial avatars for the intervention prototypes that they deemed locally appropriate to deliver messages in the study languages. Consilient Talk then used the prototype scripts and generated a series of videos of avatars relaying each message.

### Design Workshops

The IBIS study team and Busara worked together to design and plan HCD workshops in Kenya and Uganda. The goal of these workshops was to obtain feedback and insights from HCWs and community members at high risk of HIV in order to adapt the prototypes to be locally relevant, assess acceptability and initial impressions of the prototypes, and gauge whether the prototype interventions were received as intended by their theoretical underpinning in behavioral economics. We organized workshops in the same regions (Nyanza province in Kenya and southwestern Uganda) where the trial will take place. Workshops were held over two days in each country.

### Recruitment and Consenting

Recruitment took place in Kenya from November 26, 2024 to December 3, 2024, and in Uganda from January 14, 2025 to February 4, 2025. The IBIS study teams in Kenya and Uganda identified HCWs at local clinics to participate in the workshops. The team then worked with the HCWs to conduct purposive sampling and recruitment of community members. Inclusion criteria for HCWs included working as a clinical officer, nurse or doctor, and delivering HTS at a Ministry of Health (MoH) health facility in the study region. The inclusion criteria for community members included being ≥18 years of age, self-reporting HIV-negative status, and having an increased risk of HIV infection (defined as self-reporting any of the following in the past three months: having more than one sexual partner, having a known sexual partner living with HIV, having been diagnosed with a sexually transmitted infection, or having paid or received gifts or money in exchange for sex). All workshop participants provided written informed consent. Participants were given 2,000 Kenya shillings (KSh) or 55,000 Uganda shillings (UGX) (about US $15) for one day of workshop participation as reimbursement for transportation costs and compensation for their time.

### Facilitators and Staff

The workshops were led by Busara facilitators, who trained participating study staff to assist in workshop activities and guide focus group discussions. In each breakout group, there was a staff member trained in qualitative methods and focus group discussion implementation. Study staff conducted brief surveys to record basic demographic characteristics of participants. They also audio recorded focus groups to capture feedback. Trained qualitative researchers took notes and asked probing questions throughout the focus groups. Some study staff also filled out an implementation measures case report form to capture key observations, energy and participation levels, and other notable moments. At the end of each workshop day, the IBIS team gathered all study staff and facilitators to record impressions, questions, and reflections.

### Workshop Activities

The group was divided into 5 focus groups (with the exception of the Kenya workshop’s HCW, who were split into 3 focus groups) consisting of 3-8 participants to provide feedback on 2-4 intervention prototypes.

#### Focus Group Initial Discussion

Participants were asked questions by facilitators to gain a sense of their knowledge and beliefs about HIV retesting, who they considered trusted messengers and reliable sources of information about HIV testing, and what information they wanted HIV post-test counselors at health facilities to provide.

#### Prototype feedback

The facilitators played videos of the avatar delivering the prototype message for the group, and read the SMS messages aloud. Then, they asked participants a series of questions designed to elicit feedback on different aspects of the prototype (see Figure 1). The categories of these questions were as follows:

1. Initial thoughts
2. Perceptions of avatar
3. Effectiveness of video messages
4. Video design & impact on HIV retesting rates
5. Feedback on SMS follow-up messages

**Figure 1.**
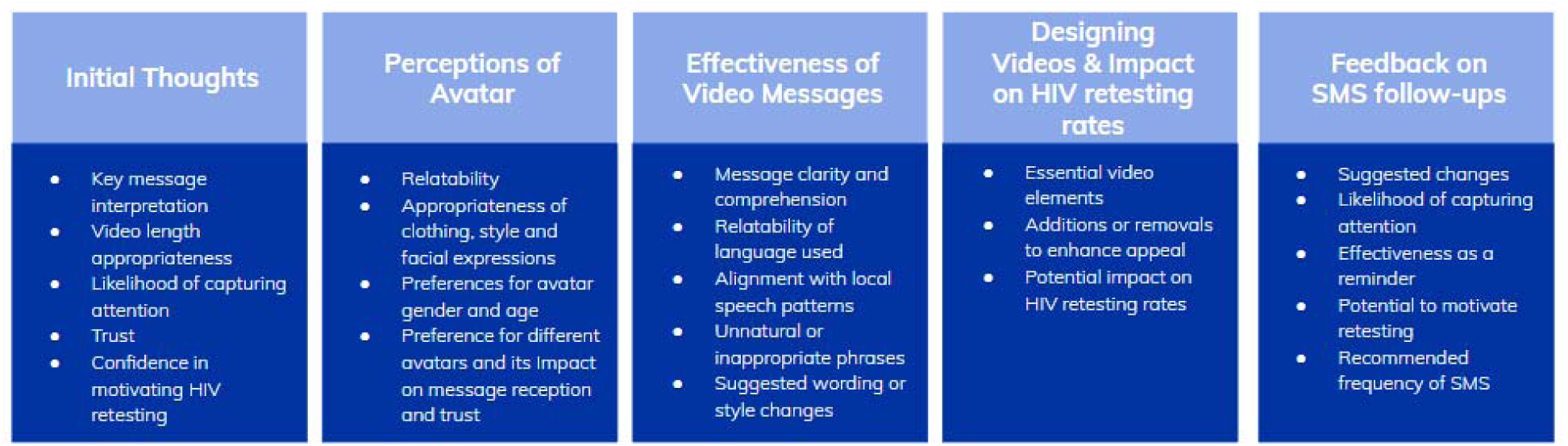
Objectives and key areas for prototype feedback.

Facilitators and staff notetakers probed as needed. They repeated this process for each prototype the focus group reviewed.

“I like, I wish, I wonder…”:

For the final portion of the workshop, participants returned to the full group. In preparation for this activity, facilitators taped up posters throughout the space. Each poster had one intervention script and was divided into four quadrants: 1) “I like,” or which parts of the script should be kept, 2) “I wish,” or which part of the script could be improved or modified, 3) “I wonder,” or what might we add to the script, and, 4) “I would remove” for language the participants wished to remove. Participants wrote their feedback on sticky notes, which they placed directly onto the relevant quadrant on the poster. Unlike the FGDs, the intention of this activity was to gather rapid feedback on many interventions from all workshop participants.

### Post-workshop analysis

After the workshops, Busara recorded and summarized the data that they had gathered from the demographic surveys, audio recordings, notes, and sticky notes. IBIS study staff recorded and summarized the information from the implementation measure forms and end of day discussion notes. The Busara team consolidated the information for each country’s workshops into two comprehensive reports with summaries, outputs, and suggested next steps and updated prototypes. With this information, the IBIS team made final updates to the prototype scripts, SMS reminders, and avatars.

## Results

Out of 36 experts contacted for intervention ideas, 15 experts submitted 23 preliminary intervention ideas that incorporated insights from behavioral science. Upon assessing the feasibility of these ideas, consolidating overlapping and repetitive ideas, and holding a vote among study investigators, we chose 11 intervention ideas to prototype. Over two day workshops, local healthcare workers and community members reacted to the 11 prototypes and offered their insights. In Homa Bay County, Kenya, workshops took place in December 2024.

Day one consisted of 16 healthcare workers, and day two consisted of 37 community members. In Uganda, workshops took place in February 2025 in Mbarara, Uganda. Day one included 16 healthcare workers, and day two held 36 community members.

Workshop participants were aged 20-57 years (Table 1). HCWs held jobs such as doctors, nurses, clinical officers, and counselors at HTS sites. Community member participants’ jobs included commercial sex work, business, and fishing trade work. Among Kenyan participants, the average age was 35 years and 28 (53%) were female. Of the 37 community members who participated, 36 (97%) had received an HIV test within the past 12 months, and all owned a mobile phone. In Uganda, there were a total of 52 workshop participants: 16 HCW and 36 community members. Among Ugandan participants, the average age was 32 years, 30 (58%) were female, and 50 (96%) owned a mobile phone. Of the 36 community members who participated, 29 (81%) had received an HIV test within the past 12 months.

**Table 1.** Participant demographic data.

| **Variable** | **Kenya** | **Uganda** | **Total** |
| --- | --- | --- | --- |
|  | **N=53** | **N=52** | **N=105** |
| Participant category |  |  |  |
| Health care worker (HCW) | 16 (30) | 16 (31) | 32 (30) |
| Community member | 37 (70) | 36 (69) | 73 (70) |
| Sex |  |  |  |
| Female | 28 (53) | 30 (58) | 58 (55) |
| Male | 25 (47) | 22 (42) | 47 (45) |
| Age, median (Q1,Q3) | 34 (29,40) | 29 (27,38) |  |
| Highest level of education |  |  |  |
| None (no formal schooling) | 0 (0) | 1 (2) | 1 (1) |
| Any Primary school | 7 (13) | 5 (10) | 12 (11) |
| Any Secondary school | 13 (25) | 23 (44) | 36 (34) |
| Post-secondary school | 33 (62) | 23 (44) | 56 (53) |
| Occupation |  |  |  |
| Business | 16 (30) | 7 (13) | 23 (22) |
| Transport | 5 (9) | 1 (2) | 6 (6) |
| Fishing | 6 (11) | 0 (0) | 6 (6) |
| Bar/pub worker | 1 (2) | 1 (2) | 2 (2) |
| Healthcare worker | 16 (30) | 21 (40) | 37 (35) |
| Farmer | 2 (4) | 0 (0) | 2 (2) |
| Sex worker | 2 (4) | 13 (25) | 15 (14) |
| Student | 1 (2) | 0 (0) | 1 (1) |
| Other | 4 (8) | 9 (17) | 13 (12) |
| **Among community members** | **N=37** | **N=36** | **N=73** |
| Has tested for HIV in last 12 months | 36 (97) | 29 (81) | 65 (89) |
| Owns a mobile phone | 37 (100) | 35 (97) | 72 (99) |
| Has shared mobile phone with others in last 12 months | 33 (89) | 24 (67) | 57 (78) |

**Data are n (%) unless otherwise specified.**

Workshop discussions yielded feedback on various aspects of the content and delivery of messages. Select quotes from workshop participants are included below to illustrate the feedback.

### HIV Retesting Knowledge

HCWs had a strong understanding of HIV retesting guidelines and the importance of frequent HIV testing among persons at increased HIV risk for early diagnosis and reducing spread of HIV. Community members were generally aware of the importance of testing frequently for HIV among high-risk persons, but many lacked awareness of the specific reasons for retesting or what groups or behaviors were considered high-risk. Both groups viewed healthcare workers and community health workers as trusted messengers.

### Message Feedback (See Table 2 for full messages)

Workshop participants felt that each video should begin with a greeting and introduction, including mention of who (i.e., what organization) was delivering the message. They also suggested that the messages reassure clients that their HIV testing results would be confidential, give information about the location of testing sites and any associated costs, and provide a specific date when a person should come for retesting instead of simply indicating that they should come within the next 3-6 months. Participants also identified words that they did not feel were appropriately translated and offered alternatives. There were several instances in which they believed that a message was judgmental or threatening towards clients. For example, participants perceived the sentence “think how disappointed your community will be if you let them down by not testing” (in the Community Benefits arm; see Table 2) as coercive, and even suggestive of threatening the client’s confidentiality.

**Table 2.**
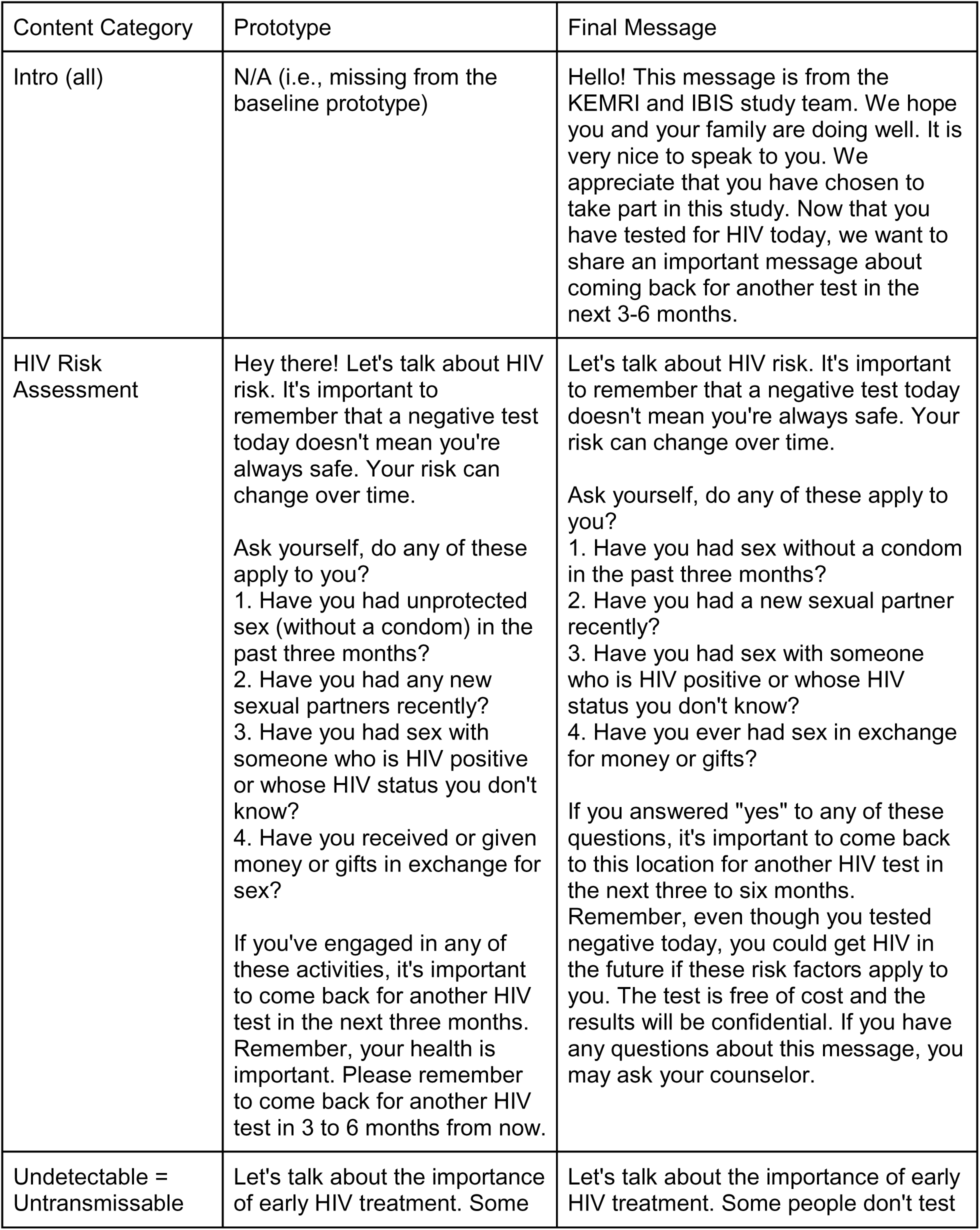
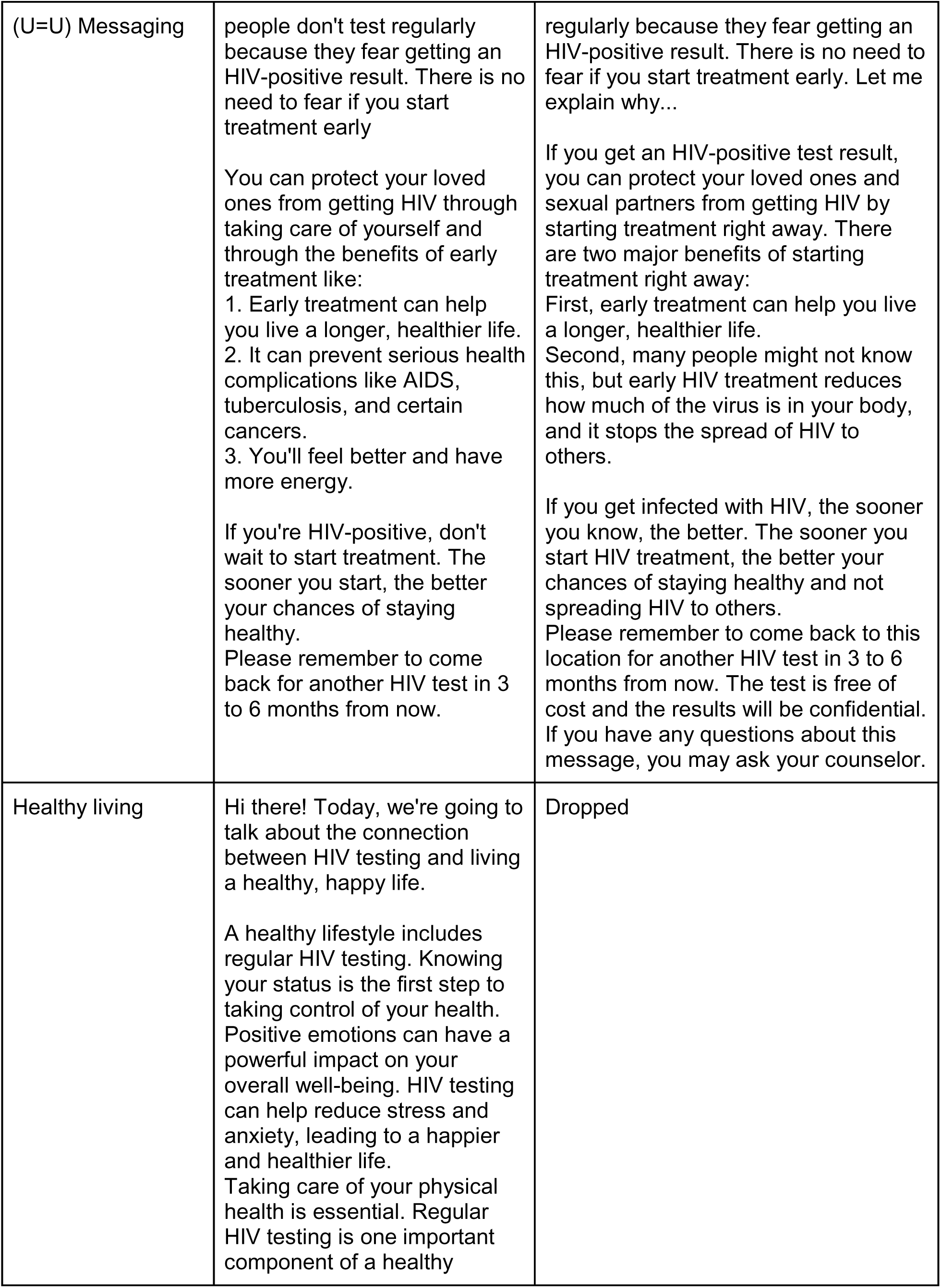
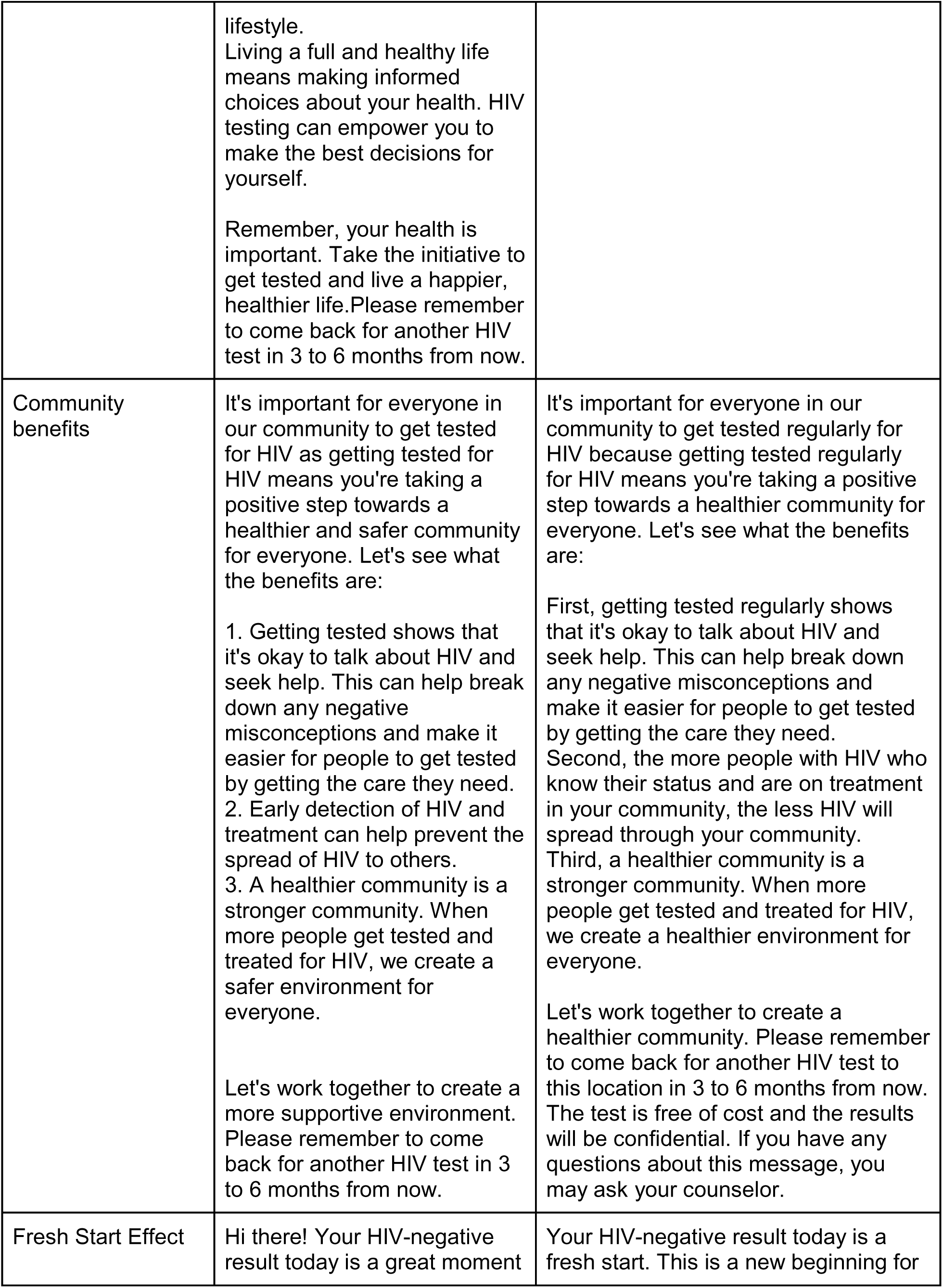
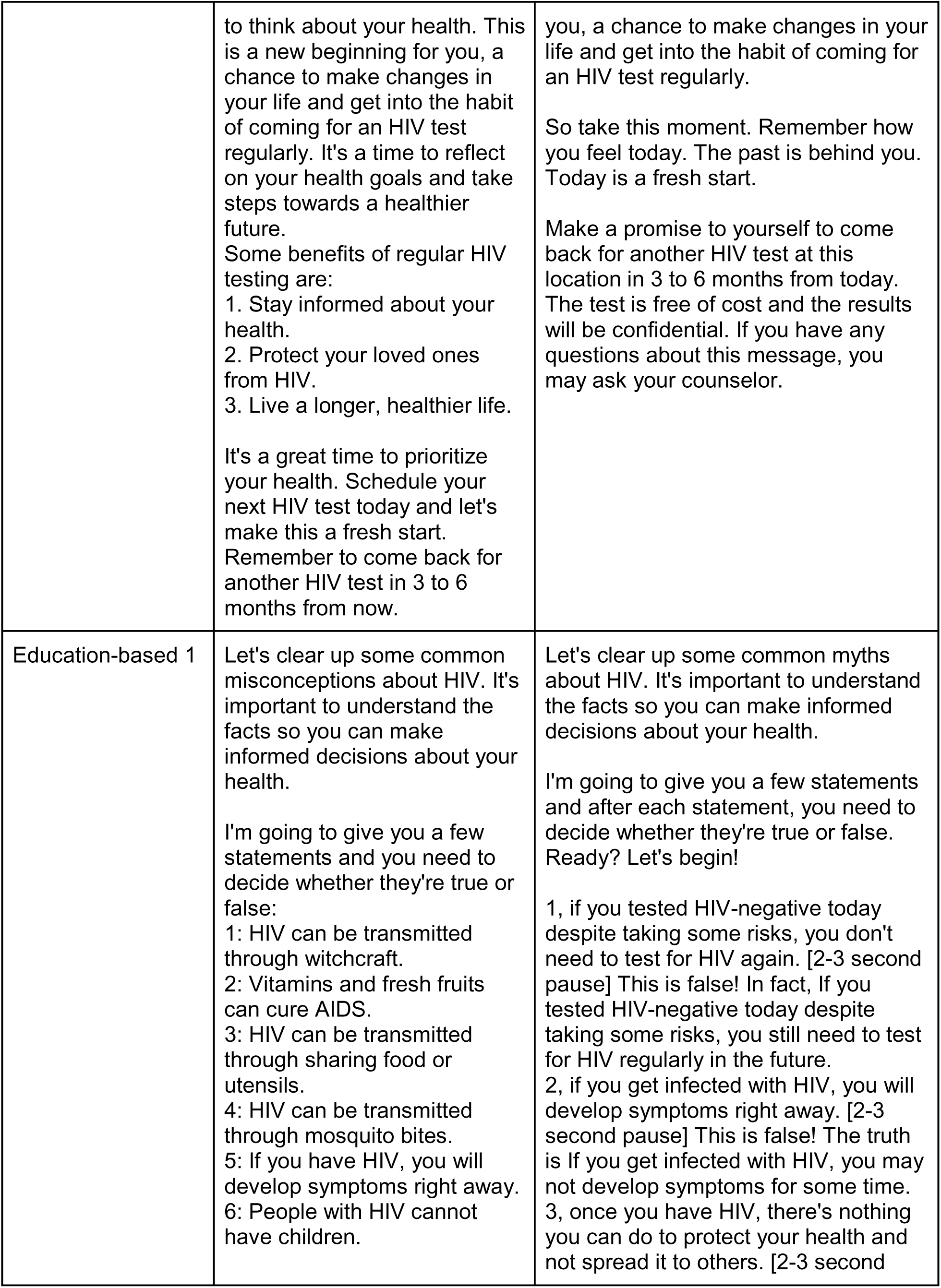
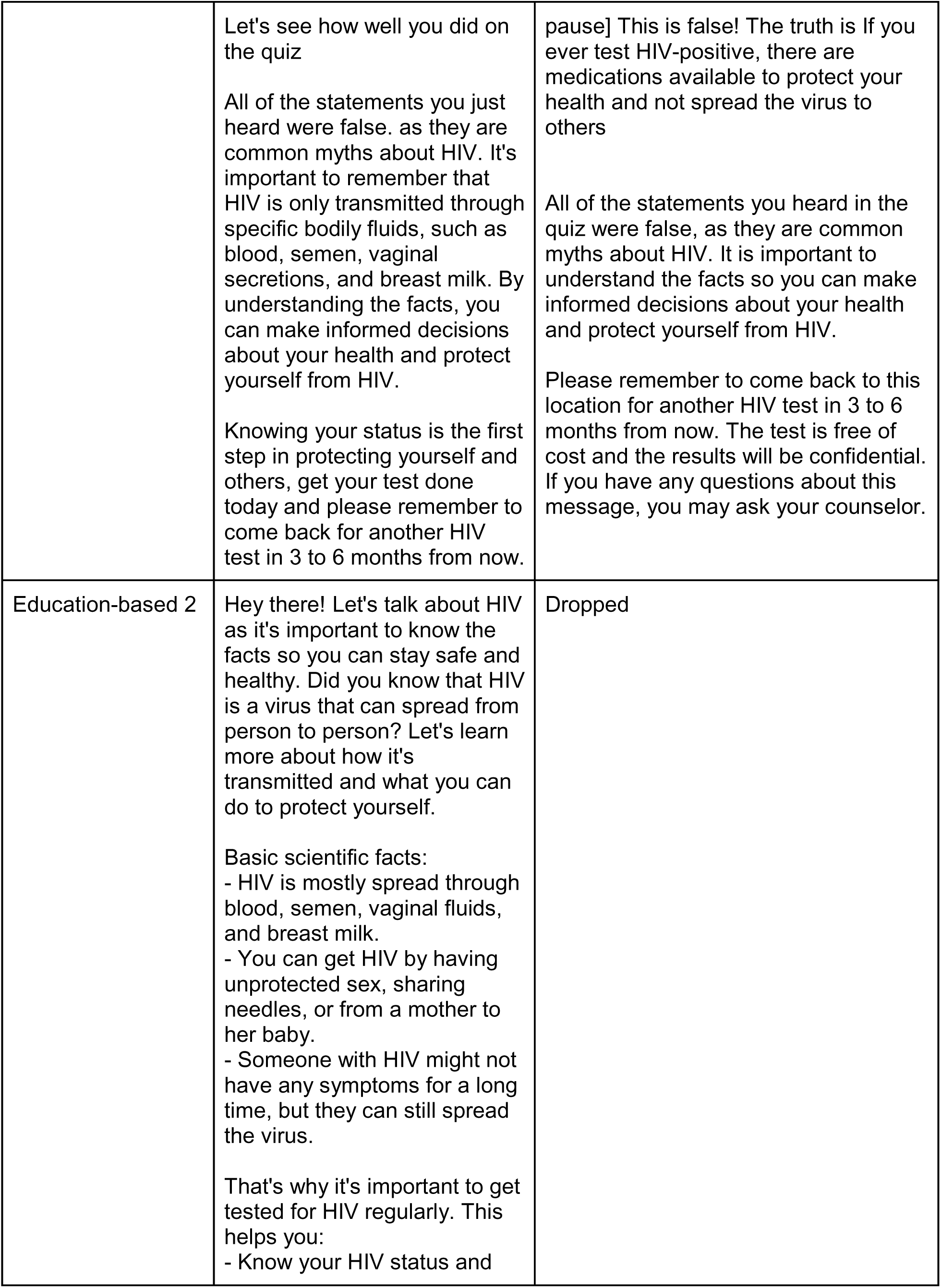
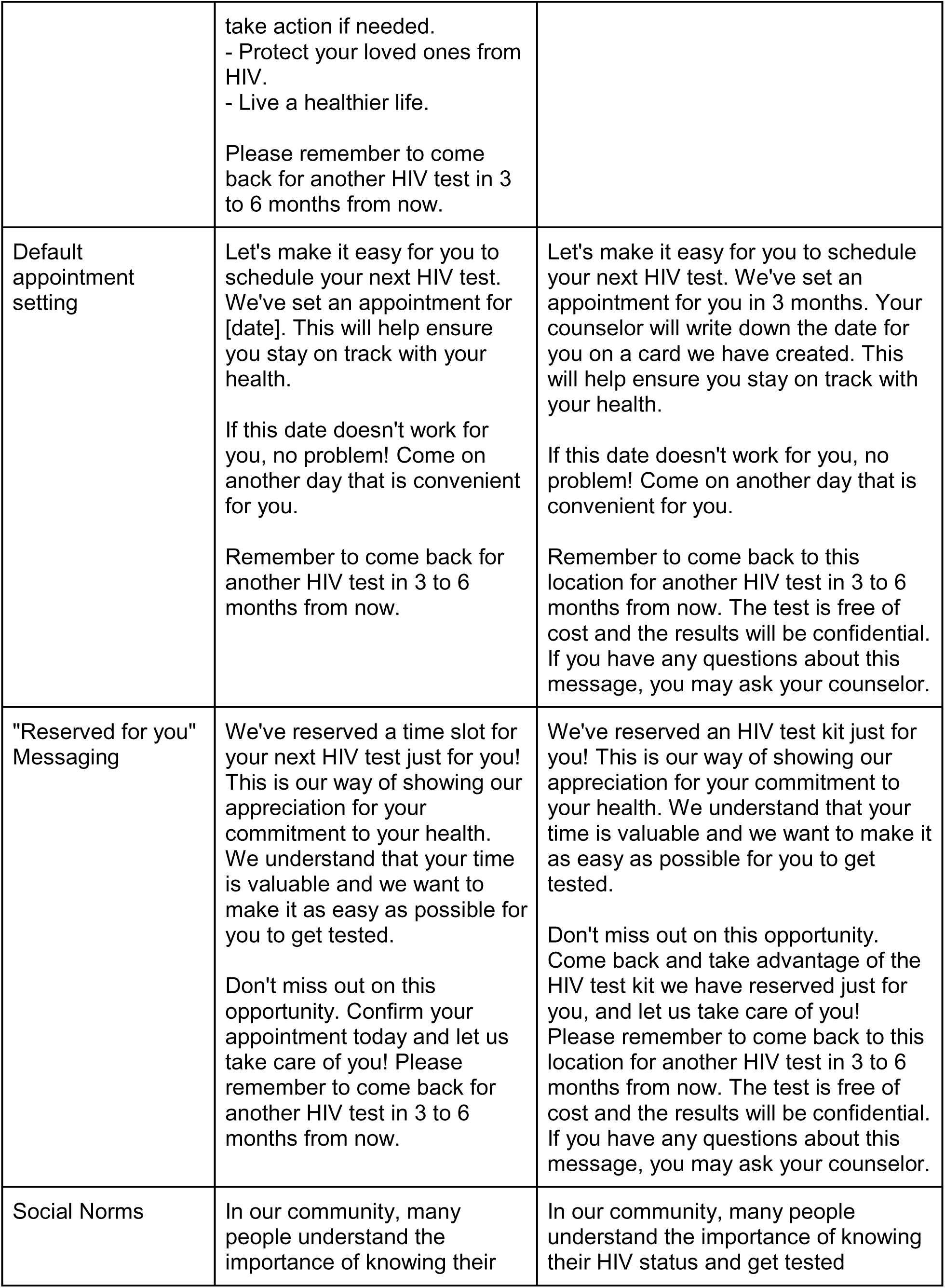
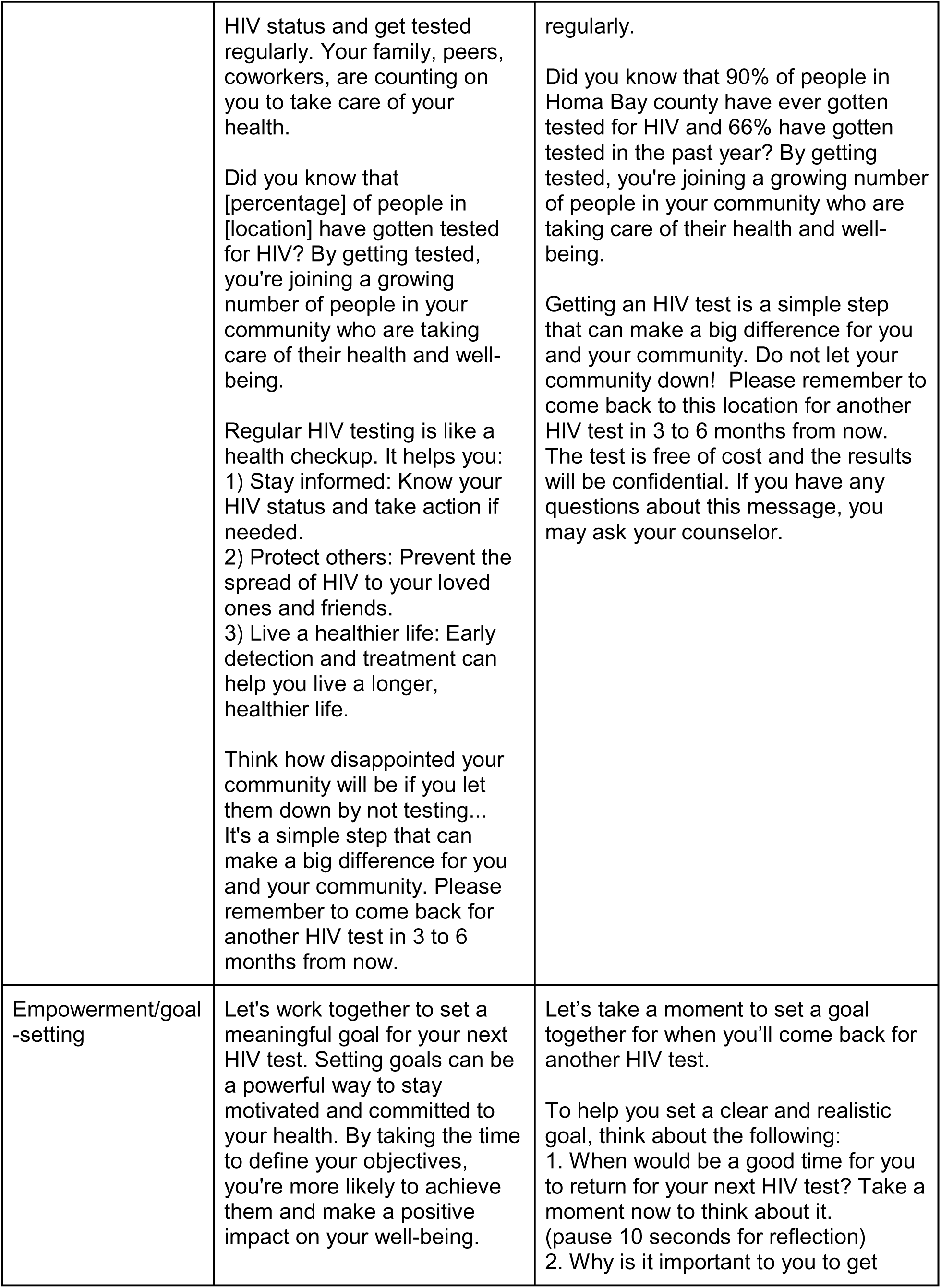
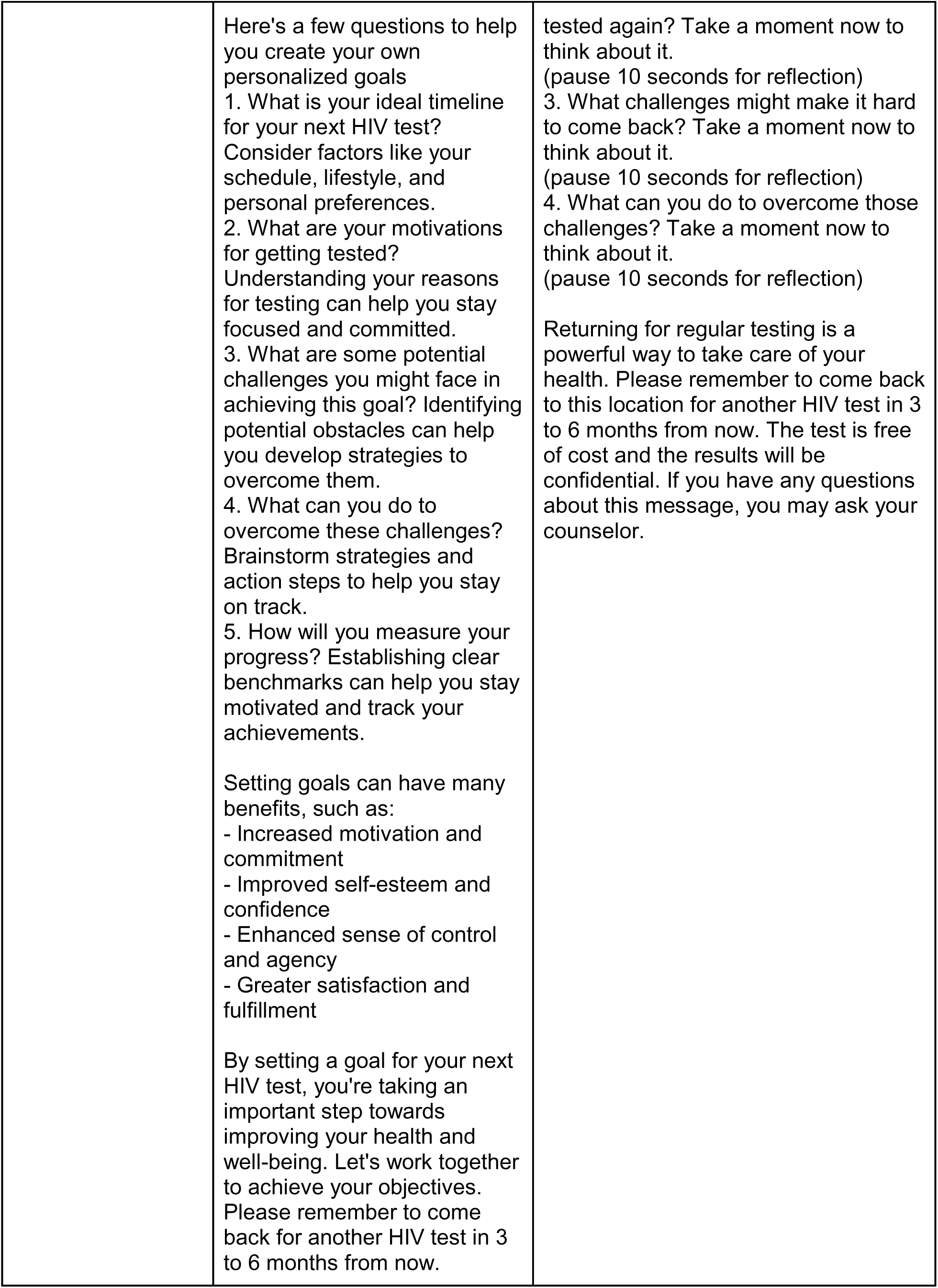
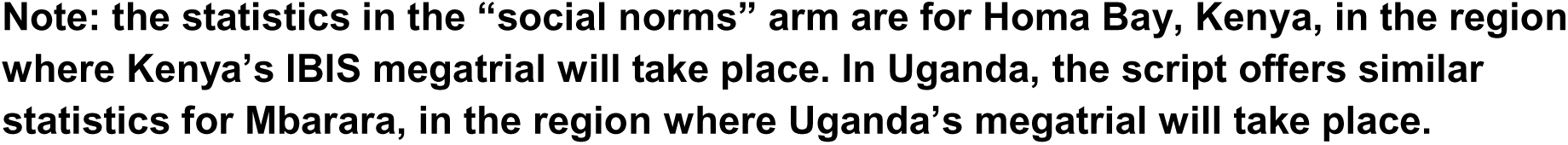
Initial “prototype” retesting messages encouraging HIV retesting, and final intervention messages post-HCD workshop feedback.

“*The part that said that I should imagine how other people will feel when I don’t get retested can be interpreted in a way that the health workers told some people that I was at the facility to test for HIV; but everything about HIV testing and retesting should be confidential.*”

-HCW, Uganda

Both community members and HCWs generally understood the underlying behavioral science concepts for each message, even if some language offended or did not resonate with them.

They referenced specific phrases in the message that conveyed the concepts, such as “we have set aside a date for your HIV retest” and “don’t let this opportunity pass you” in the “‘Reserved for You’ Messaging” arm, and “[HIV] transmission can take place without signs and symptoms” in the “Education Based 2” arm (see Table 2). Notable exceptions included the “Social Norms” message, which many participants saw as more threatening than compelling, and the “Education Based 1” arm, which some Ugandan participants interpreted as perpetuating HIV myths, rather than debunking them.

“*The quote ‘living a good life’ is really inspiring, for you to be in a healthy life or a good life, you need to know your HIV status.”*

-HCW, Kenya

### AI Avatar feedback

Uganda workshop participants found the prototype intervention avatars to be rigid and unengaging. They suggested using avatars that incorporated body language and hand gestures, and adding visuals to capture viewers’ attention. Kenya workshop participants had concerns about the clothing some female avatars wore (i.e., clothing considered too revealing), and recommended more culturally-appropriate clothing for the avatars. In particular, there was one female avatar which the Kenya group strongly disliked, as they believed she looked too thin and sickly, as if she had HIV, which the group believed might perpetuate HIV stigma (Figure 2). Participants across workshops suggested depicting the avatars as trusted community members, especially medical professionals, by including a hospital background or dressing the avatar in a white coat.

**Figure 2.**
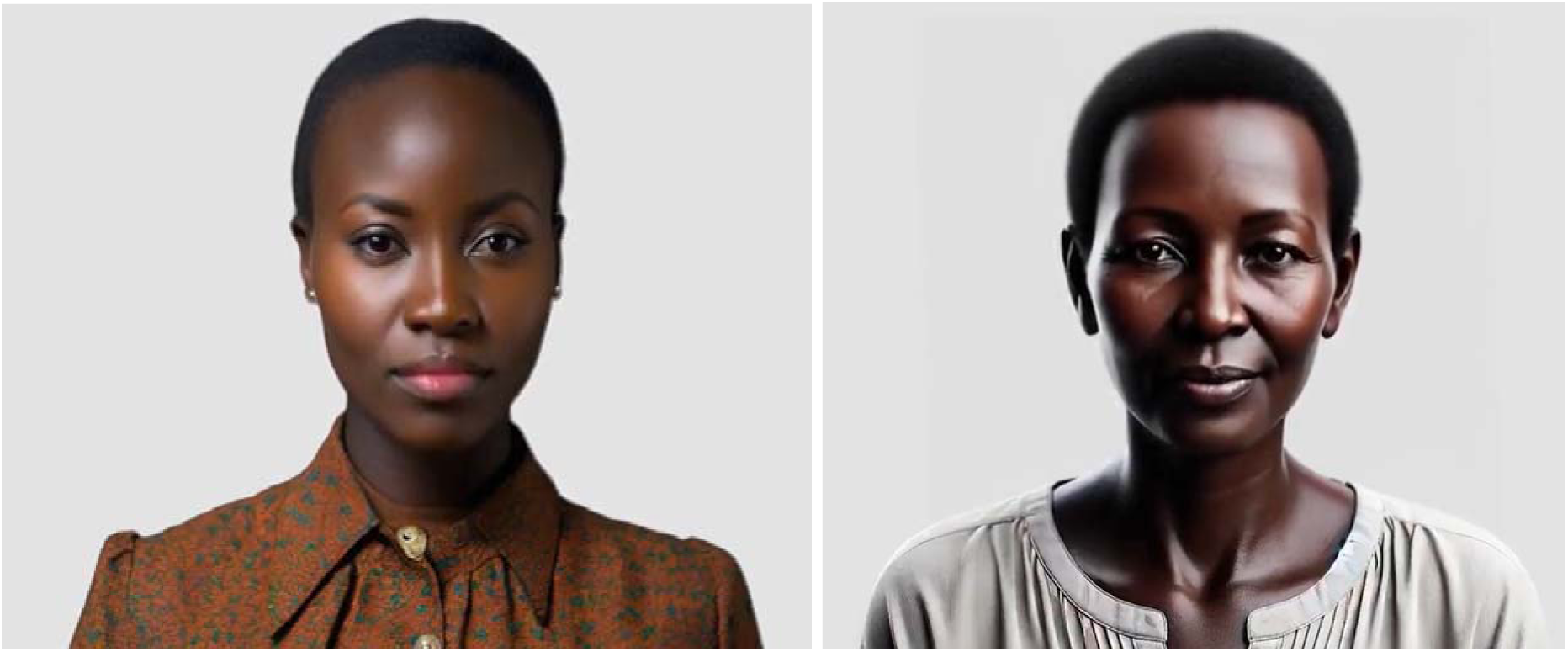
Cynthia, Uganda avatar (left) whom participants deemed acceptable and Ruby, Kenya avatar (right), whom participants deemed unacceptable

“*To make it appealing I would add some visual illustrations to the video. I don’t think the person in the video should stand still as if he is singing the national anthem.*”

-HCW, Uganda

*“She is dressed like a TV presenter. In Africa, if you go to a drug shop it should have a nurse in uniform but now this person has this information but cannot be identified as a health worker.”*

-Community member, Uganda

### SMS feedback

Participants suggested that the identity of the sender be made clearer, in order to emphasize that a credible messenger (such as the study team) had sent them. They also advocated for language which kept the message concise and easily readable, and which clearly stated details about local testing sites, hours of operation, and the fact that HIV testing is free of charge at these government-supported health centers. Focus group members were particularly wary about language which mentioned or alluded to HIV, in case a partner or family member had access to the phone. Instead, they requested more neutral language which maintained client privacy.

*“These SMS messages did not capture my attention because SMS should be short and these ones are too long. I don’t think anyone has the time to read those long SMS.”*

-HCW, Uganda

“*For the part that states ‘Remember when we talked about setting a goal for your next HIV test?’ Suppose I used my spouse’s phone number or he gets to find such a message from my phone and I did not inform him that I was going for a HIV test and they get to see this kind of message – it will bring a lot of issues…*”

-Community Member, Kenya

### Workshop participant observation

Facilitators and staff from the Kenya HCW workshops, the first of the workshops, noted that participants tended to start off very engaged in workshop activities, and then grew less focused as the day progressed. These groups reviewed 3-4 prototypes, and their workshop ran later than expected due to delays in arrivals. In an effort to limit attention fatigue, we decided to limit the number of prototypes a focus group reviewed to 2-3. We also asked participants to come earlier in order to start on time. In the subsequent workshops in Kenya and Uganda, there remained some issues with lack of concentration throughout the day, but it was not as prominent. Notes from the Kenya HCW workshops also indicated that the “I like, I wish…” activity was difficult to conduct in a small physical space, so we made an effort to spread out the space for implementation of this activity during the remaining workshops. Overall, staff reported that participants were excited to participate in the FGDs and offer their feedback to help shape the messages.

### Final prototypes (See Table 2)

With workshop feedback, we added a uniform greeting, with an introduction and ending to each message. We modified the language in the messages according to message-specific feedback, and identified the source of that message. In the SMS, we removed language which referenced HIV directly, replacing it with language that did not explicitly mention “HIV”. We also made arm-specific modifications, such as changing language in places where participants felt it poorly translated or inappropriate for the audience.

There were some places where we opted not to include participant feedback. Largely, this was in cases in which participant feedback resulted in intervention overlap, given plans for the future randomized controlled megastudy trial to test effects of distinct messaging interventions. We also limited modifications which weakened the BE theory underlying an intervention.

Furthermore, some of the suggested avatar modifications (e.g. changing the background, the avatar’s tone of speech, adding hand gestures, etc.) were not feasible for Consilient Talk to incorporate.

## Discussion

Over the course of two human-centered design workshops in rural Kenya and Uganda, community members and healthcare workers provided extensive feedback on behavioral economics-informed video avatar and follow-up SMS messages designed to encourage HIV retesting among individuals at high risk of HIV. The workshops offered opportunities to iteratively adapt the interventions for local acceptability and contextual relevance. Embedding trained qualitative research assistants within workshop groups and conducting participant observation allowed for the identification of challenges and content-specific concerns.

The HCD workshops provided a structured mechanism for integrating community perspectives into the intervention. Participants had strong reactions to certain language and imagery, such as the perception of threat in “social norms” messages, discomfort with references to HIV in SMS content, and disapproval of some avatars’ appearance. We made various changes to address this feedback, including adding a uniform greeting and ending to each message, identifying the source of that message, and making intervention-specific language modifications. We removed language in the SMS messages that directly referenced HIV to address privacy concerns, and adjusted inaccurate translations and terms. These findings highlight the importance of integrating local community members and experts into the design phase, and thus ensuring that content deemed acceptable by researchers will be equally acceptable to the target audience.

We also obtained important insights on the acceptability of AI-generated avatars for delivering health promotion messages, which will be useful as such tools become more commonplace in the future. The availability of AI-generated avatars as a design feature is a recent innovation, and one which allows for the rapid development and iterative modification of customized messages delivered in local languages by a local-appearing messenger that can be changed over time. In contrast to videos recorded with human actors, the avatar videos are easier to modify through an iterative process. However, little is known about how people receive these messages and the suggestions they have about avatar usage. This study provides some general lessons in this area. For one, workshop participants provided detailed suggestions on how to modify the avatar messages and messengers to a local context. The avatars attracted attention and commentary - some participants found the avatars rigid (i.e., lacking in body language) - but many participants found them to be professional and persuasive messengers. Though not all modifications to the avatars could be incorporated (such as the addition of hand gestures) with the avatars we selected, future AI-generated avatars are likely to allow for a range of modifications and HCD workshops offer an opportunity to better customize messengers according to local preferences. Furthermore, many stylistic preferences, such as gender and clothing preferences of avatars, were straightforward to address and indeed led to changes in the interventions, emphasizing the importance of community input on messenger identity to ensure the messenger appealed to a local audience and did not generate unintended, negative perceptions.

Incorporating participant insights into the content and design of health promoting messages can preemptively address challenges that may arise during the trial that will test the effectiveness of these messages. Comparable approaches in other HIV prevention contexts have shown that collaborative design processes not only refine intervention content but also foster a sense of ownership among participants, potentially improving uptake and adherence (30). By addressing these concerns early in the design process, rather than post-implementation, we reduced the likelihood of disengagement, offense, or unintended harms during the trial.

Not all of the insights gleaned during the workshop led to prototype alterations, nor did it encompass all areas of the study in which we wished to gain feedback. For example, a key goal of the workshops was to assess whether each prototype conveyed a message that invoked its underlying behavioral theory. In each focus group, there was overall information about whether the group understood the message, what it was trying to convey about retesting, and how it conveyed that message. However, it is apparent that participant feedback tended to pertain largely to more stylistic elements of the video such as the avatar’s appearance and visual elements of the video, as well as language considerations such as the translations used and terminology. This feedback is deeply important in ensuring that the videos will resonate with the intended audience and effectively communicate the retesting message. However, these decisions may have distracted participants from thinking about the underlying behavioral concepts, which are central to the study.

Some of the suggested stylistic and language changes - or suggestions to include elements across interventions - also diluted the behavioral theory underlying particular arms, or blurred the language that distinguished between key elements of the arms. In these instances, we had to make decisions to maintain the core principle of each message. This sometimes entailed not including feedback in the final interventions, or finding new language which addressed feedback without weakening the message. In a scoping review of Human-Centered Design applications in global health, Bazzano et al speak to the need for “tolerance for ambiguity” in design thinking, which can be difficult to employ in empirical research processes of public health (31). This may be true to some degree in an intervention such as the IBIS megatrial, which went into the workshops with a focused aim and prototypes to be fine-tuned by participants, rather than completely regenerated.

Our study has several limitations. First, workshops were held in Kenya’s Nyanza province and southwestern Uganda. As such, the feedback from this workshop may not be broadly generalizable beyond the study regions. In addition, participants may have been subject to social desirability bias, and refrained from expressing negative feedback to discussion group leaders. Others may not have felt comfortable speaking up in a group setting, especially community members who were reflecting on potentially sensitive subject areas of HIV and sexual behavior. Finally, participants pointed out several mistranslations of messages throughout discussions. In some cases, this may have impacted participants’ understanding of the message.

## Conclusion

Human centered design workshops in Kenya and Uganda generated valuable, context-specific insights that shaped the final avatars, scripts, and messages encouraging HIV retesting for the IBIS megatrial. This study demonstrates the critical role of early, iterative community engagement in optimizing interventions that seek to promote changes in health behavior. Next steps include the implementation of these messages in a large-scale trial in which we will assess their effectiveness in promoting HIV retesting.

## Data Availability

The data presented in the manuscript is based on qualitative methods. Excerpts of the transcripts relevant to the study are included throughout the paper

## Acknowledgements

Ethical approval for study activities was granted by the University of California San Francisco Human Research Protection Program, the Makerere University School of Medicine Research and Ethics Committee, and Kenya Medical Research Institute Scientific and Ethics Review Unit.

This research was supported by the National Institute of Mental Health (NIMH) [R01MH132438] and the National Institute on Alcohol Abuse and Alcoholism (NIAAA) [K24AA031211]. The funder played no role in study design, data collection, analysis and interpretation of data, or the writing of this manuscript.

## Statement on AI use

During the preparation of this work the authors used ChatGPT in order to refine wording and ensure clarity in certain sections. After using this tool/service, the authors reviewed and edited the content as needed and take full responsibility for the content of the published article.

## References

1. Banerjee AV, Duflo E, Glennerster R, Kothari D. Improving immunisation coverage in rural India: clustered randomised controlled evaluation of immunisation campaigns with and without incentives. BMJ. 2010 May 17;340(may17 1):c2220–c2220. doi:10.1136/bmj.c2220

2. Brune L, Giné X, Goldberg J, Yang D. Facilitating Savings for Agriculture: Field Experimental Evidence from Malawi. Economic Development and Cultural Change. 2016 Jan;64(2):187–220. doi:10.1086/684014

3. Giné X, Karlan D, Zinman J. Put Your Money Where Your Butt Is: A Commitment Contract for Smoking Cessation. American Economic Journal: Applied Economics. 2010 Oct 1;2(4):213–35. doi:10.1257/app.2.4.213

4. Jensen R. Do Labor Market Opportunities Affect Young Women’s Work and Family Decisions? Experimental Evidence from India *. The Quarterly Journal of Economics. 2012 May;127(2):753–92. doi:10.1093/qje/qjs002

5. Karlan D, McConnell M, Mullainathan S, Zinman J. Getting to the Top of Mind: How Reminders Increase Saving. Management Science. 2016 Dec;62(12):3393–411. doi:10.1287/mnsc.2015.2296

6. Milkman KL, Patel MS, Gandhi L, Graci HN, Gromet DM, Ho H, et al. A megastudy of text-based nudges encouraging patients to get vaccinated at an upcoming doctor’s appointment. Proc Natl Acad Sci USA. 2021 May 18;118(20):e2101165118. doi:10.1073/pnas.2101165118

7. PATH. Human-centered design [Internet]. [cited 2026 Jan 19]. Available from: https://www.path.org/what-we-do/health-systems-strengthening/human-centered-design/

8. Cheney C. Devex [Internet]. 2016 [cited 2026 Jan 19]. A human-centered approach to design for development. Available from: https://www.devex.com/news/sponsored/a-human-centered-approach-to-design-for-development-87978

9. Holeman I, Kane D. Human-centered design for global health equity. Information Technology for Development. 2020 Jul 2;26(3):477–505. doi:10.1080/02681102.2019.1667289

10. Smith PJ, Joseph Davey DL, Schmucker L, Bruns C, Bekker LG, Medina-Marino A, et al. Participatory Prototyping of a Tailored Undetectable Equals Untransmittable Message to Increase HIV Testing Among Men in Western Cape, South Africa. AIDS Patient Care and STDs. 2021 Nov 1;35(11):428–34. doi:10.1089/apc.2021.0101

11. Baumann SE, Rabin MA, Devkota B, Hawk M, Upadhyaya K, Shrestha GR, et al. Centering Communities in Global Health: Using Human-Centered Design to Facilitate Collaboration and Intervention Development. Community Health Equity Research & Policy. 2025 Jan;45(2):167–85. doi:10.1177/2752535X241264331

12. World Health Organization. Consolidated guidelines on differentiated HIV testing services. Geneva: World Health Organization; 2024. 1 p.

13. Giguère K, Eaton JW, Marsh K, Johnson LF, Johnson CC, Ehui E, et al. Trends in knowledge of HIV status and efficiency of HIV testing services in sub-Saharan Africa, 2000–20: a modelling study using survey and HIV testing programme data. The Lancet HIV. 2021 May;8(5):e284–93. doi:10.1016/S2352-3018(20)30315-5

14. Marson K, Ndyabakira A, Kwarisiima D, Camlin CS, Kamya MR, Havlir D, et al. HIV retesting and risk behaviors among high-risk, HIV-uninfected adults in Uganda. AIDS Care. 2021 May 4;33(5):675–81. doi:10.1080/09540121.2020.1842319

15. Cawley C, Wringe A, Isingo R, Mtenga B, Clark B, Marston M, et al. Low Rates of Repeat HIV Testing Despite Increased Availability of Antiretroviral Therapy in Rural Tanzania: Findings from 2003–2010. Tang JW, editor. PLoS ONE. 2013 Apr 23;8(4):e62212. doi:10.1371/journal.pone.0062212

16. Kranzer K, Van Schaik N, Karmue U, Middelkoop K, Sebastian E, Lawn SD, et al. High Prevalence of Self-Reported Undiagnosed HIV despite High Coverage of HIV Testing: A Cross-Sectional Population Based Sero-Survey in South Africa. Landay A, editor. PLoS ONE. 2011 Sep 28;6(9):e25244. doi:10.1371/journal.pone.0025244

17. Regan S, Losina E, Chetty S, Giddy J, Walensky RP, Ross D, et al. Factors Associated with Self-Reported Repeat HIV Testing after a Negative Result in Durban, South Africa. Caylà JA, editor. PLoS ONE. 2013 Apr 23;8(4):e62362. doi:10.1371/journal.pone.0062362

18. Moyo E, Moyo P, Murewanhema G, Mhango M, Chitungo I, Dzinamarira T. Key populations and Sub-Saharan Africa’s HIV response. Front Public Health. 2023 May 16;11:1079990. doi:10.3389/fpubh.2023.1079990

19. Zeleke EA, Stephens JH, Gesesew HA, Gello BM, Ziersch A. Acceptability and use of HIV self-testing among young people in sub-Saharan Africa: a mixed methods systematic review. BMC Prim Care. 2024 Oct 15;25(1):369. doi:10.1186/s12875-024-02612-0

20. Maughan-Brown B, Venkataramani AS. Accuracy and determinants of perceived HIV risk among young women in South Africa. BMC Public Health. 2018 Dec;18(1):42. doi:10.1186/s12889-017-4593-0

21. Muravha T, Hoffmann CJ, Botha C, Maruma W, Charalambous S, Chetty-Makkan CM. Exploring perceptions of low risk behaviour and drivers to test for HIV among South African youth. Marotta C, editor. PLoS ONE. 2021 Jan 22;16(1):e0245542. doi:10.1371/journal.pone.0245542

22. Orne-Gliemann J, Zuma T, Chikovore J, Gillespie N, Grant M, Iwuji C, et al. Community perceptions of repeat HIV-testing: experiences of the ANRS 12249 Treatment as Prevention trial in rural South Africa. AIDS Care. 2016 Jun 2;28(sup3):14–23. doi:10.1080/09540121.2016.1164805

23. Nangendo J, Katahoire AR, Armstrong-Hough M, Kabami J, Obeng-Amoako GO, Muwema M, et al. Prevalence, associated factors and perspectives of HIV testing among men in Uganda. Francis JM, editor. PLoS ONE. 2020 Aug 7;15(8):e0237402. doi:10.1371/journal.pone.0237402

24. Walker GR. Emotive Media as a Counterbalance to AIDS Messaging Fatigue in South Africa: Responses to an HIV/AIDS Awareness Music Video. AIDS Education and Prevention. 2022 Feb;34(1):17–32. doi:10.1521/aeap.2022.34.1.17

25. Paschen-Wolff MM, Restar A, Gandhi AD, Serafino S, Sandfort T. A Systematic Review of Interventions that Promote Frequent HIV Testing. AIDS Behav. 2019 Apr;23(4):860–74. doi:10.1007/s10461-019-02414-x

26. Busara. Busara [Internet]. Busara; 2026 [cited 2026 Jan 19]. More about Busara. Available from: https://busara.global/more-about-busara/

27. Consilient. Consilient [Internet]. Consilient Research; 2026 [cited 2026 Jan 19]. Thematic Areas. Available from: https://consilientresearch.org/thematic-areas/

28. MIT SOLVE. SOLVE [Internet]. MIT SOLVE; 2024. 2024 Global Health Equity Challenge: Consilient Talk. Available from: https://solve.mit.edu/solutions/86905

29. Consilient Talk. Consilient Talk [Internet]. 2026 [cited 2026 Jan 19]. Consilient Talk. Available from: https://consilient-talk.org/

30. Leung CL, Naert M, Andama B, Dong R, Edelman D, Horowitz C, et al. Human-centered design as a guide to intervention planning for non-communicable diseases: the BIGPIC study from Western Kenya. BMC Health Serv Res. 2020 Dec;20(1). doi:10.1186/s12913-020-05199-1

31. Bazzano AN, Martin J, Hicks E, Faughnan M, Murphy L. Human-centred design in global health: A scoping review of applications and contexts. Virgili G, editor. PLoS ONE. 2017 Nov 1;12(11):e0186744. doi:10.1371/journal.pone.0186744

